# The King’s College London Coronavirus Health and Experiences of Colleagues at King’s Study (KCL CHECK) protocol paper: a platform for study of the effects of coronavirus pandemic on staff and postgraduate students

**DOI:** 10.1101/2020.06.16.20132456

**Authors:** Katrina A.S. Davis, Sharon A.M. Stevelink, Ammar Al-Chalabi, Gabriella Bergin-Cartwright, Rupa Bhundia, Howard Burdett, Ewan Carr, Grace Lavelle, Daniel Leightley, Candice Middleton, Timothy Nicholson, Lucy O’Neill, Catherine Polling, Aoife Ruane, Alice Wickersham, Matthew Hotopf, Reza Razavi

## Abstract

**Introduction:** We will use an occupational sample to study the impact of COVID-19 on current staff and postgraduate research students at a large UK university. The cohort study will address some of the key questions about COVID-19 for the international community, while also providing feedback to the employer and educator.

**Methods and analysis:** Participants were recruited via email to their University email address. Administrative records were available to compare the composition of volunteer participants to underlying staff and postgraduate student populations of the University. The study comprises a baseline survey, longitudinal follow-up surveys and a viral antibody study. Baseline information was collected in April 2020 including demographics, working situation, current stresses and worries, mental health and neurological symptoms. Personal experiences of COVID-19, indirect experiences and attitudes towards the pandemic were queried, as well as satisfaction with communication and support at work. Longitudinal surveys will assess changes in COVID-19 exposure and mental health. A viral antibody detection component is being planned and will also be longitudinal in nature.

**Ethics and dissemination:** Ethical approval has been gained from KCL’s Psychiatry, Nursing and Midwifery Research Ethics Committee (HR-19/20-18247). Participants were provided with information and agreed to a series of consent statements before enrolment. Data are kept on secure servers with access to personally identifiable information limited. Researchers may apply to have access to pseudonymised data. Findings will be disseminated internally to the University and participants, and externally through scientific publications.

## Background

### Context

The COVID-19 pandemic is a global threat that may have biological, behavioural, emotional, and social impact. Key unknowns include the longer-term health effect of infection with SARS-CoV-2 (the virus that causes COVID-19), the impact of blanket physical isolation, and the immune response to SARS-CoV-2. There are many policy-relevant questions for the national and international health response, such as around immune status testing and the need for interventions to tackle loneliness during physical isolation. There are also relevant questions on a smaller scale for employers and educators about how best to support staff and students through a threat that is social and economic, as well as to health.

To answer these questions, we need flexible and rapid methods of data collection. Studies that engage with participants longitudinally will also be preferred, to observe changes in clinical state, coping strategies, and behaviours through what may be a likely prolonged period of disruption. Web-based platforms for recruitment and data-collection (for example^1-3^) are adaptable, scalable at low cost and can provide near real-time feedback on population data. Web-based data collection also avoids face-to-face contact between researchers and participants, which minimises the risk during the pandemic. However, many web-based studies of COVID-19 have used convenience samples recruited via social media.^4^ These introduce inherent biases and are ill-equipped to determine prevalence of mental disorders. There is a need for research where the characteristics of the population being sampled from are known.

The staff and postgraduate research (PGR) student population at King’s College London, a large Russell Group University, provides a valuable population to study. Administrative records provide detailed information on the composition of the staff and PGR student populations, making it possible to evaluate the generalisability of respondents. Cohort studies involving staff and PGR students can inform both high-level scientific questions and small-scale decisions regarding how the University communicates and supports people during the pandemic.

### Current knowledge

The combination of the new international biological threat and the social measures put in place to manage the pandemic is unprecedented, and therefore without historical data. A recently published paper by mental health science experts - research leaders, funders and people with lived experience - lays out some of the biggest unknowns and research priorities arising from COVID-19.^5^ They suggest the need to collect high-quality data in two areas: (i) the direct effect of SARS-CoV-2 on those infected and (ii) the adverse effects on population mental health of the COVID-19 pandemic and public health responses.

Regarding the infection itself, there is concern over its effect on brain function and mental health. SARS-CoV-2 can affect the brain,^6 7^ although it is not currently clear whether this is due to the action of the virus itself, the immune response, or the stress of the illness and treatment. Severe infection (that necessitating hospital admission) is probably more commonly associated with overt brain dysfunction,^8^ but there have not yet been evaluations of brain dysfunction after mild infections - and therefore there is little idea of the population-level effect.

At the same time, wellbeing may suffer due to the sudden changes in society from public health measures. Literature about quarantine^9^ combined with data collected during early stages of ‘lockdown’ due to COVID-19^10,12^ suggest that, even without personal or indirect exposure to COVID-19, this social change can cause increased distress. It also appears that the risk factors for distress may be different than for COVID-19 itself. For instance, although older people may be more vulnerable to direct effects of SARS-CoV-2, they appear more resilient than younger and working-age people to the negative psychological effects of the social and economic changes it has caused.^13^ Levels of anxiety and depression may rise and life satisfaction fall due to isolation, economic hardship and lack of autonomy, as well as practical difficulties, frustration and increased use of alcohol. Data on who is most at risk of mental health problems as a result of COVID-19, and through what mechanisms may contribute to whole-population mitigation efforts.^14^ Data needs to be longitudinal and may need to be supplemented by qualitative research or novel data collection techniques.

The KCL-CHECK research platform will address some of the unanswered questions about the impact of the COVID-19 pandemic, and what organisations such as universities can do to minimise the impact on staff. It will be able to investigate the direct effect of what we anticipate will mainly be mild manifestations of COVID-19 by repeated assessment with validated mental health, neurological and immunological measures over up to 18 months of follow-up. We will also collect indirect experiences of the illness and the difficulties arising from the pandemic response, especially those stresses related to work, to look at the wider effects of COVID-19.

### Research aims and hypotheses

- To study the impact of the short- medium- and long-term COVID-19 pandemic on health and wellbeing outcomes among KCL staff and PGR students
- To identify unmet needs among KCL staff and PGR students related to the pandemic response at KCL as they emerge

In particular, we aim to study the following (with *key hypotheses* identified):

1. Determine the psychological and psychiatric impacts (including hazardous / harmful alcohol use) of the wider COVID-19 pandemic,

- Individuals reporting more psychological impact related to COVID-19 (worry, preoccupation, etc.) will experience worse psychiatric outcomes, and the relationship will be stable over time
- Reported stressors, including physical social isolation, during the pandemic will be associated with worse psychiatric outcomes, and this will happen in a cumulative pattern (the more stressors over time, the worse the outcome)
2. Identify psychosocial indicators of resilience and vulnerability

- People with characteristics sometimes associated with social disadvantage (female gender, junior employment, younger working age, non-White British ethnicity, disability and living with children) will experience greater indirect impact of the COVID-19 pandemic (such as childcare and other caring responsibilities, social isolation and other stressors)
- Job insecurity among staff and worries about education among PGR students will be longitudinally associated with poorer psychiatric outcomes and reduced wellbeing
3. Identify individuals with COVID-19 and follow them up with respect to psychological impact, mental health, and neurological health; using the remainder of the cohort to act as control group

- Experiencing COVID-19 symptoms (and once known, immunological SARS-CoV-2 infection status) will be associated with worse psychiatric outcomes or more neurological symptoms
- People who think they have had COVID-19 (and once known, immunological SARSCoV-2 infection status) will have less general anxiety and worry of contracting COVID-19, along with engaging less with physical isolation and other government advice.
4. To understand the level of underlying infection in the cohort using home antibody testing and to track immunity if possible. Exploratory analyses:

- Examine the association and discrepancies between whether participants think they have had COVID-19 and/or reported COVID-19 symptoms with immunological SARSCoV-2 infection status, in particular, examining for potentially asymptomatic SARSCoV-2 infection, and describing the profile of these if found.
- Track consecutive immunological SARS-CoV-2 tests (both IgG and IgM) to look at any variation, which may represent changes in immunity.

The research platform that we have created would also enable different research studies to build on this, such as recruiting people for intervention studies.

## Methods and analysis

### Study protocol

All King’s College London (KCL) employees (n≈9800) and PGR students (n≈2500) were eligible to take part. Participants were recruited initially via email sent from the principal investigators to staff and PGR students to their University email addresses. Recruitment was also encouraged via internal communications.

There are three study modules that recruited via the platform:

i. Baseline survey
ii. Longitudinal surveys
iii. Viral antibody testing

The initial recruitment period ran from April 16^th^ to 30^th^ 2020. Follow-up is planned for an initial period of 18 months. For any increase in study length, ethical approval will be sought. The projected study timeline is shown in Table 1.

**Table 1.** Projected timeframe for research platform

|  | Timepoint (month) |  |  |  |  |  |  |  |  |  |  |  |
| --- | --- | --- | --- | --- | --- | --- | --- | --- | --- | --- | --- | --- |
| Data collection | 1 (1) | 2 (1) | 3 (2) | 4 (2) | 5 (3) | 6 (3) | 7 (4) | 8 (4) | ... | 34 (17) | 35 (18) | 36 (18) |
| Demographics | X |  |  |  |  |  |  |  |  |  |  |  |
| Measures of:<br>COVID-19 symptoms<br>Depression<br>Anxiety<br>Loneliness | X | X | X | X | X | X | X | X | every fortnight | X | X | X |
| Work challenges<br>Social distancing<br>Indirect experience of COVID-19<br>Indirect impact of COVID-19<br>Psychometrics and attitude<br>Wellbeing<br>Psychiatric<br>Alcohol<br>Neurological | X |  |  |  | X |  |  |  | every two months |  | X |  |
| KCL-specific measures | X |  | X |  | X |  | X |  | as needed | X |  |  |
| Virology |  |  |  |  |  | X |  |  | up to every two months |  | X |  |

Potential participants were able to view the participant information sheet and could also email the research team via a dedicated study email address to ask any questions prior to taking part. Data for the surveys are collected via Qualtrics, an online survey platform. Registration started online with an informed consent process through a series of prompts on the website. When signing up to take part, participants were asked permission for follow-up through (i) a follow-up survey every two months, and (ii) a brief fortnightly survey. If they consented to follow-up, they were then asked about participating in the viral antibody module (meaning a testing kit would be posted to their address), being contacted for future studies, and receiving SMS text notifications. No incentives were given for participation.

The baseline survey was launched before the viral antibody testing, so that validated testing kits could be obtained. Pilot testing of the viral antibody module will take place in June 2020, with roll-out shortly after. The results from the viral antibody testing and from the follow-up questionnaires will be linked to the baseline survey by a unique ID number which was generated automatically.

#### Eligibility criteria for platform and baseline survey

Included participants must be:

1. A current KCL staff member or PGR student
2. Aged 18 and over at time of participation
3. Able and willing to give informed consent to participate in the survey
4. Able to understand and communicate in written English and use a web browser

#### Eligibility for longitudinal surveys

Participants of the longitudinal surveys must be participating in the baseline survey, and additionally be:

1. Willing to provide an email address to receive follow-up surveys

#### Eligibility criteria for optional viral antibody testing

Participants of the viral antibody testing module must be participating in the baseline and longitudinal surveys, and additionally:

1. Be willing and able to conduct home antibody testing with a supplied rapid Immunoassay Test Cassette
2. Supply a UK address for delivery of the testing kits
3. Agree to informing the research team of the results of the testing kit

#### Participant withdrawal

Participants are free to withdraw from further surveys and modules at any time by emailing the dedicated study email address. We aim to publish non-identifiable data in brief summaries within weeks of data collection, and we explain in the participant information that this means that if participants withdraw their consent, we cannot withdraw their data from completed analyses or ongoing analyses where the data has already been extracted and pseudonymised. We will withdraw their data from the dataset for future analyses if requested.

## Measures

### Baseline survey

As shown in Table 1 and supplementary materials, the baseline survey collected information on: demographics and occupation; lifetime physical and mental health; exposure to COVID-19 at work and home and risk factors for COVID-19. Impacts of the COVID-19 pandemic on the participants, their family and close networks, and on wider society were collected. A symptom checklist adapted from a COVID-19 symptom tracker app^15^ was used. A bespoke selection of measures previously validated for use in the general population provided the psychiatric (including Patient Health Questionnaire-PHQ9^16^ and Generalised Anxiety Disorder-GAD7^17^), psychological, and neurological outcomes (see supplementary materials for details). This detailed questionnaire will be repeated (apart from demographics and lifetime health) every two months.

### Longitudinal surveys

i. Every fortnight those participants who agreed to fortnightly surveys will receive a link via email to a very brief survey (max 15 minutes), including COVID-19 symptoms^15^, symptoms of anxiety (GAD7) and depression (PHQ9), loneliness and change in drinking habits. It may sometimes also contain selected topical questions, particularly those of relevance to KCL, such as thoughts about returning to work.
ii. Every two months, participants will receive a detailed questionnaire as above.

### Viral antibody testing

A Rapid Immunoassay Test Cassette will be used to measure evidence of previous infection of SARS-CoV-2 based on the presence of IgM and IgG antibodies to the ‘spike’ protein. Subject to a successful pilot, the test kit used will be the SureScreen Diagnostics COVID-19 IgG/IgM Rapid Test Cassette, and the procedure will be that cassettes are sent to the consenting participants’ home address, along with a lancet for providing a blood spot and the necessary reagents, as well as a piece of paper with a printed rectangle and their study ID number. Participants will be asked to place a drop of blood and a drop of reagent onto the cassette, wait ten minutes, and take a photograph of the cassette lying inside the rectangle on the paper provided, with ID number visible. Participants will then be able to upload this via a website, which can then be matched to their data using the ID number by a designated researcher. Viral antibody testing will be repeated on up to eight occasions approximately two months apart during the study to track exposure and immunity over time.

Home testing with Rapid Immunoglobulin Test Cassettes have the advantages of being convenient for participants, rapid, and low cost. Validation has shown the IgG portion of the test has the potential to be accurate, with proprietary testing of the IgG portion showing 97% sensitivity, 99% specificity (38 SARS-CoV-2 positive and 143 negative cases)^18^ and independent testing at St Thomas’s Hospital showing 96% sensitivity, 100% specificity (29 samples from SARS-CoV-2 positive cases 20-30 days after initial symptoms and 50 negative samples)^19^. However, home testing may not be as ideal a situation as in the laboratory, which will be monitored during the trial. At the time of the launch of the study, SureScreen and similar Rapid Immunoassay Tests were not approved for clinical use, and it will be made clear to participants that this is not a clinical test.

## Statistical methods

Response rates will be calculated based on administrative records from KCL on the numbers of staff and PGR students (from the human resources department and the Higher Education Statistics Agency, HESA^20^). Information from the baseline survey will be summarised with appropriate summary statistics (e.g. mean and SD for continuous measures; frequencies and proportions for categorical). Results will be presented overall as well as stratified by staff/PGR student. All scales will be scored according to published instructions. Missing items will be handled according to these instructions or mean imputed (when <10% of items are missing) where no instructions are available.

The extent of missing information will be summarised by item. Differences in the composition of survey respondents compared to the target populations (all KCL staff and PGR students) will be summarised. This will draw on administrative records for KCL staff (on age, gender, ethnicity, pay grade, role (academic, clinical, professional services, etc.), working hours (full-time/part-time), and contract type. Information on the composition of PGR students in terms of age, gender, and ethnicity will be derived from HESA data^20^. Weights will be derived to adjust for differences in sample composition using post-stratification and raking. Weights will be applied to all analyses and weighted percentages will be provided in addition to raw counts.

The prevalence of probable mental disorders, neurological and physical health outcomes will be summarised using descriptive statistics. Associations between COVID-19 experiences and mental health will be tested using generalised linear models, with adjustments for measured confounders. Longitudinal data will be modelled using mixed effect generalised linear models, with random intercepts and slopes for individual participants (as appropriate), to allow comparison of individual trajectories (e.g. in mental health) overtime.

We will explore the specific hypotheses listed above and we have pre-defined key constructs. “Psychiatric outcomes” (mentioned as an outcome in some hypotheses) are defined as shown in box 1. “Coronavirus symptoms” (mentioned as an exposure in some hypotheses) will be defined in two algorithms, one with high sensitivity, and the other with relatively high specificity. The former is “one of fever, persistent cough or loss of smell/taste”, which became the symptoms requiring action according to the UK Department of Health and Social Care on 18 May 2020; the latter is the algorithm derived by researchers using data from the symptom tracking app, shown in box 2, which has a positive predictive value of 0.69 and negative predictive value of 0.75 in the tested sub-sample of the app population.^21^

*Free text* Free text will be analysed using two main methods: i) doing a qualitative framework analysis to identify and classify themes in the data and ii) using natural language processing (NLP) to identify apparent groups in the data. These two methods will then be compared to enable decision making about whether N LP could be used as the principal form of future analyses either a) using the same agnostic algorithm if it has performed comparably to the framework analysis or b) by creating a rules based algorithm based on the findings of the framework analysis.

*Viral Antibody testing:* Using repeated testing on the same participant using the same brand of Rapid Immunoglobulin Test Cassette will not only allow us to confirm infection, but also study the timing of seroconversion from IgM to IgG and the timing of the loss of IgM and possibility of loss of IgG in the cohort over time.

## Data management and oversight

The study is registered on the King’s Data Protection Register (ref: DPRF-19/20-14373). Dr Sharon Stevelink is the designated data controller.

Online survey responses are collected using Qualtrics survey software via the King’s College London Qualtrics account. Data is stored on servers located in the UK. Access to identified data is restricted to individuals managing the survey. Data required for analysis will be pseudonymised by separation of any identifying information and stored on KCL OneDrive. Any interactions with Qualtrics are logged and will be audited by a designated researcher. All researchers in the team have completed data protection training. Requests to access the data is subject to application to the principal investigators with a valid research proposal that is regarding the COVID-19 pandemic and is in accordance with the UK Policy Framework for Health and Social Care research. Where the applicant is outside of KCL, a data-sharing agreement will be drawn up by the KCL legal team. Since this survey included free text and there was the possibility that participants had identified themselves or their line managers, before providing free text to researchers, free text will briefly be reviewed by one of the researchers who is managing the survey, and identifying information will be removed.

It is important to note that for participant recruitment and the respect of their confidentiality within the KCL organisation it was stated that “those in authority in your employing organisations will not have access to your data”. Since two of the Principal Investigators are senior KCL managers, they will not have access to individual level data.

### Viral antibody testing

The data collected as part of the viral antibody testing will consist of the presence or absence of IgM and/or IgG antibodies based on photographs of the Rapid Immunoassay Test Cassette supplied by participants. Images will be received via a secure pathway and associated with the participant dataset by a designated researcher. For participants having difficulties taking a picture and sending this to the research team, alternative arrangements will be made.

Individual test data will not be shared with healthcare professionals or other organisations, and this will be emphasised to participants.

## Data storage and security

Data will be collected over 18 months and processed in accordance with the General Data Protection Regulations on the basis of public interest. A data processing agreement has been signed with Qualtrics who will be acting as the data processor. Data will be retained for four years after the start of the study to allow for analyses and scientific publication. After four years data not needed for current research purposes will be securely destroyed.

## Ethical and safety considerations

This study has been approved by KCL’s Psychiatry, Nursing and Midwifery Research Ethics Committee (HR-19/20-18247).

### Confidentiality

One feature of this study is that participants will be asked to provide feedback on King’s College London’s handling of the COVID-19 situation, and this will be fed back to key figures in the University to inform the approaches to support the health and wellbeing of staff and PGR students during the COVID-19 pandemic. It is therefore vital that no identifiable information will be available in this feedback to maintain the confidentiality of the participants. As such, it was decided we would collect information on where people worked at an faculty level, not departmental, and not to collect details of the funder for PGR students (only internal/external) to avoid inadvertent unmasking of participants. Full text responses that identified individuals will also be removed prior to research.

### Participant safety

The nature of the survey will be clearly explained, and participants are able to stop participating at any time or skip questions. Signposting information will be provided both after questions regarding suicidal ideation and at the end of the survey signposting people to support services.

The result of the antibody assay will not be escalated to a GP or local testing service; potentially this could cause a delay in receiving appropriate advice or treatment. The viral antibody test is not licensed for clinical use, but there is also a risk that participants interpret the test results as health information or as a sign of clinical immunity. It will be made clear to the participant that if the test comes back negative, it does not necessarily mean that the participant has not been infected by SARS-CoV-2, nor does a positive test mean that people are no longer susceptible to getting the infection in the future. Participants will be encouraged to follow national and NHS guidelines, seeking medical advice in the normal way if they have any concern about their wellbeing.

### Participant burden

The baseline survey including the consent procedure took approximately 40 minutes to complete. It is anticipated that the 2-monthly follow-up online surveys can be completed in 30 minutes. The fortnightly surveys take only 5-15 minutes to complete (depending on occasional topical questions). Participants can complete surveys at a time convenient for them and can take breaks and come back to the survey at any time in the next seven days. Participants can also withdraw from future iterations of the study at any time.

The viral antibody testing will be performed by the participant at their home address. This reduces the participant burden of travelling to a research location and removes the need for face-to-face contact, therefore minimising risk of contact with SARS-CoV-2, but does push some of the technical burden onto participants. A set of instructions will be provided with test kits including guidance on how to produce and utilise a blood spot, and how to share the test outcome with the research team.

## Dissemination

Findings from each survey round will be rapidly assembled to give timely feedback at stakeholder meetings (i.e. for KCL managers) using a mixture of formal and informal internal reports. The findings will inform the KCL strategy on managing and supporting staff and PGR students during the current COVID-19 pandemic, and how this is communicated. Other internal reports will be disseminated, including periodic reports of findings aimed at the platform participants.

Further research and analysis will be prepared for academic publication. Some publications will require the full longitudinal data, but others will be planned on cross-sectional or limited longitudinal data. We will aim to place publications on pre-print servers at the point of submission to research journals, to reach an audience more quickly than would otherwise be the case in this rapidly evolving situation. There has been increased use of pre-print servers to disseminate and identify research regarding COVID-19.^22 23^

## Funding and roles

KCL-CHECK is funded by King’s College London. Additionally, this paper represents independent research part-funded by the National Institute for Health Research (NIHR) Biomedical Research Centre at South London and Maudsley NHS Foundation Trust and King’s College London.

All investigators are employed by King’s College London. Two of the principal investigators are also senior managers in King’s College London. Decisions regarding study design, and the collection, and management of data were made by the named investigators. Decisions regarding the analysis, interpretation and writing of reports will be carried out by the named investigators.

The investigators have regular meetings to agree study design and analysis decisions. The publication, prior to analysis, of this protocol with key hypotheses has been planned to contribute to transparency in future academic publications. The decision to submit reports to publication will remain under the ultimate authority of the principal investigators. The views expressed in this and future papers will be those of the authors and not necessarily those of the King’s College London, the NHS, the NIHR or the Department of Health and Social Care

## Conclusions

KCL-CHECK is a web-based research platform to study the impact of COVID-19 on a specific occupational sample of current staff and postgraduate research students at a large UK university. The planned research comprises a baseline survey, longitudinal follow-up surveys and a viral antibody study. One aim is to be able to give close to real-time feedback to KCL managers, which may be able to enhance the wellbeing of the participants and their colleagues. As a scientific study of the effect of COVID-19 pandemic on health and wellbeing, concentrating on psychiatric and neurological outcomes, it has strengths due to the recruitment from a population where the underlying characteristics are known, rapid roll-out, use of validated measures for outcomes, and explicit hypotheses being agreed prior to analysis of results. The study is planned to last 18 months, although some analyses on cross-sectional and interim results will be available before the study end.

## Data Availability

There is no associated data.

## Acknowledgements

This paper represents independent research part-funded by the National Institute for Health Research (NIHR) Biomedical Research Centre at South London and Maudsley NHS Foundation Trust and King’s College London. The views expressed are those of the author(s) and not necessarily those of the NHS, the NIHR or the Department of Health and Social Care.

Thanks to colleagues and friends who helped edit and test surveys

## Box 1.

### Proposed psychiatric outcomes

| *Box 1. Proposed psychiatric outcomes* |  |  |
| --- | --- | --- |
|  | *Cross-section* | *Longitudinal (change in -)* |
| ***Depression*** | self-reported worsening in symptoms of depression (as described in the PHQ9) | change in status/score on PHQ9 |
| ***Anxiety*** | self-reported worsening in symptoms of anxiety (as described in GAD7) | change in status/score on GAD7 |
| ***Hazardous alcohol use*** | scoring positive on Alcohol Use Disorder Identification Test (AUDIT) and self-reports increase in alcohol use | change in status/score on AUDIT |
| ***Suicidal ideation*** | suicidal ideation since the pandemic | change in status on suicidal ideation |
| ***Self-harm*** | self-harm since the pandemic | change in status on self-harm |
| *PHQ status (to be confirmed): 0-4 none, 5-9 mild, 10-14 moderate, 15-19 mod-severe, 20+ severe* |  |  |
| *GAD status (to be confirmed): 0-4 none, 5-9 mild, 10-14 mod-severe, 15+ severe* |  |  |
| *AUDIT status (to be confirmed): 0-7 none, 8-15 mild, 16-19 moderate, 20+ severe* |  |  |

## Box 2

Suggested formula for identification of COVID-19 cases from symptoms recorded in symptom tracker app^15^ from which we adapted our COVID-19 screening questions. As published.^21^

If result of following > 0.5 then regarded as positive case. If <= 0.5 then negative case -1.32 - (0.01 x age) + (0.44 x sex) + (1.75 x loss of smell and taste) + (0.31 x persistent cough) + (0.49 x severe fatigue) + (0.39 x skipped meals)

Where male participants have sex=l and female participants have sex=0, and symptoms are marked as 1 if present and 0 otherwise.

\**original formula split into >0.5 and <0.5 with no allowance for exactly 0.5*

## Notes

### Competing Interest Statement

All authors are employed by the organisation that commissioned and funded the study (King's College London)

### Clinical Trial

n/a

### Author Declarations

Ethical approval has been gained from KCL's Psychiatry, Nursing and Midwifery Research Ethics Committee (HR-19/20-18247)

### Summary of Updates

Protocol originally allowed for withdrawing participants from the study if they did not complete two 2-monthly surveys, but it was decided that since this was not in the ethics or patient information sheet that this should not be used. Therefore it has been removed from the protocol paper.

